# *FLNC* and *MYLK2* gene mutations in a Chinese family with different phenotypes of cardiomyopathy

**DOI:** 10.1101/2020.05.10.20097519

**Authors:** Xianyu Qin, Ping Li, Hui-Qi Qu, Yichuan Liu, Yu Xia, Shaoxian Chen, Yongchao Yang, Shufang Huang, Pengju Wen, Xianwu Zhou, Xiaofeng Li, Yonghua Wang, Lifeng Tian, Hakon Hakonarson, Yueheng Wu, Jian Zhuang

**Affiliations:** Guangdong Cardiovascular Institute, Guangdong Provincial Key Laboratory of South China Structural Heart Disease, Guangdong Provincial People’s Hospital, Guangzhou, Guangdong Province, China; Center for Applied Genomics, The Children’s Hospital of Philadelphia, Philadelphia, Pennsylvania, USA; Department of Pediatrics and Division of Human Genetics, University of Pennsylvania, Philadelphia, Pennsylvania, USA; Prenatal Diagnosis Center, Department of Obstetrics and Gynecology, Guangdong Provincial People’s Hospital, Guangzhou, Guangdong Province, China, China

**Keywords:** dilated cardiomyopathy, *FLNC*, hypertrophic cardiomyopathy, *MYLK2*, sudden cardiac death

## Abstract

**Background:** Mutations in the sarcomeric protein filamin C (*FLNC*) gene have been linked to hypertrophic cardiomyopathy (HCM), in which they increase the risk of ventricular arrhythmia and sudden death. In this study, we identified a novel missense mutation of *FLNC* in a Chinese family with HCM and interestingly a second novel truncating mutation of *MYLK2* in one family member with different phenotype.

**Methods:** We performed whole-exome sequencing in a Chinese family with HCM of unknown cause. To validate the function of a novel mutation of *FLNC*, we introduced the mutant and wild-type gene into AC16 cells (human cardiomyocytes), and used western blotting to analyze the expression of *FLNC* in subcellular fractions, and confocal microscope to observe the subcellular distribution of the protein.

**Results:** We identified a novel missense single nucleotide variant (*FLNC* c.G5935A [p.A1979T]) in the family, which segregates with the disease. *FLNC* expression levels were equivalent in both wild type and p.A1979T cardiomyocytes. However, expression of the mutant protein resulted in cytoplasmic protein aggregations, in contrast to wild type *FLNC*, which was distributed in the cytoplasm and did not form aggregates. Unexpectely, a second truncating mutation, NM_033118:exon8:c.G1138T:p.E380X of the *MYLK2* gene, was identified in the mother of the proband with dilated cardiomyopathy, but absent in other subjects.

**Conclusion:** We identified the *FLNC* A1979T mutation as a novel pathogenic variant associated with HCM in a Chinese family, as well as a second causal mutation in a family member with a distinct phenotype. The possibility of more than one causal mutations in cardiomyopathy warrants clinical attention, especially for patients with atypical clinical features.

## 1 Introduction

Hypertrophic cardiomyopathy (HCM) is a common cause of sudden cardiac death (SCD) in the young (1-3). Significant left ventricular hypertrophy that cannot be explained by other causes is the main clinical feature of HCM (4). It is generally believed that HCM is an autosomal dominant hereditary disease. Previous studies have identified that more than half of the HCM-related pathogenic genes encode sarcomere proteins (4). Amongst known causal HCM variants, those in the *FLNC* gene (OMIM *102565) have been reported to be associated with the high incidence of SCD in young age (5, 6). *FLNC* is mainly expressed in cardiac and skeletal muscles and is a type of actin cross-linking protein(7). It is a known gene that can cause skeletal muscle disease, mutations of which can result in myofibrillar myopathy (MFM)(7) and distal myopathy(8). Approximately one-third of MFM cases have heart involvement (7, 9). Lately, pathogenic variants in *FLNC* were reported to cause HCM, but without skeletal muscle involvement(5). Missense pathogenic variants of *FLNC* were also reported in individuals with restrictive cardiomyopathy (RCM) (10). Subsequently, some studies found that *FLNC* variants can cause dilated cardiomyopathy (DCM) and arrhythmogenic right ventricular cardiomyopathy (ARVC) (6). In general, filamin C-related HCM has a higher proportion of SCD when compared with HCM cases without *FLNC* mutations (5). However, the pathogenesis of FLNC-related cardiomyopathy is elusive, and it is essential to study the pathogenesis of *FLNC*.

In this study, we sequenced a family with HCM and SCD. We identified a novel missense variant of *FLNC* in the family. We investigated the factional effects of the novel mutation (c. G5935A [p.A1979T]) in the *FLNC* gene to better understand the pathogenic mechanisms of *FLNC* mutation in HCM. Interestingly, one family member with distinct phenotype could be explained by a second truncating mutation of the *MYLK2* gene.

## Materials and Methods

### Patients and family

Local approval for this study was provided by the Ethics Review Board of the Guangdong Provincial people’s Hospital and all participants signed informed consent forms. The patients in the present study were from the Chinese Han population. The proband (□-1), a 29-year old, asymptomatic female, was diagnosed with HCM at 20 years of age based on the published criteria(11), including electrocardiography showing left ventricular hypertrophy and echocardiography showing a hypertrophic septum (19mm), New York Heart Association (NYHA) grade I and no left-ventricular outflow obstruction (LVOTO). An electrocardiogram indicated sinus tachycardia and left ventricular hypertrophy. Her past medical history has not included any treatment to date.

She had a family history of sudden death with a maternal aunt and a maternal uncle died of SCD, both at 20 years old. Her father (□-2) is healthy. Her mother (□-1), who experienced palpitations and short of breath, was diagnosed with dilated cardiomyopathy (in contrast to HCM of the proband) and heart failure. Ultrasonic echocardiography showed dilated cardiomyopathy, significantly decreased left ventricular systolic function, severe mitral regurgitation, severe tricuspid regurgitation and severe pulmonary hypertension, which indicated left ventricular non-compaction. Electrocardiogram results of the mother revealed sinus rhythm, occasional atrial premature beats, and intraventricular block. II-2 is currently on the waiting list for a heart transplant. The proband’s maternal grandmother, who died from heart failure at 91 years of age, was diagnosed with sub-aortic stenosis at 66 years of age and HCM at 88 years. Her maternal aunt (□-3) complained of chest tightness and had a history of coronary artery disease and stent intervention, complicated with nodular goiter and gastritis. Ultrasonic echocardiography showed HCM. Her maternal uncle (□-5) did not have any apparent symptoms. His electrocardiogram suggested primary atrial ventricular block and ultrasonic echocardiography indicated mitral valve prolapse, with mild valve regurgitation and ascending aortic enlargement. There was no significant change in the results of the ultrasonic echocardiography through ten years of follow-up. The pedigree information is shown in Fig.1. All the affected subjects in this study had an isolated cardiac phenotype, without skeletal muscle involvement.

**Fig. 1.**
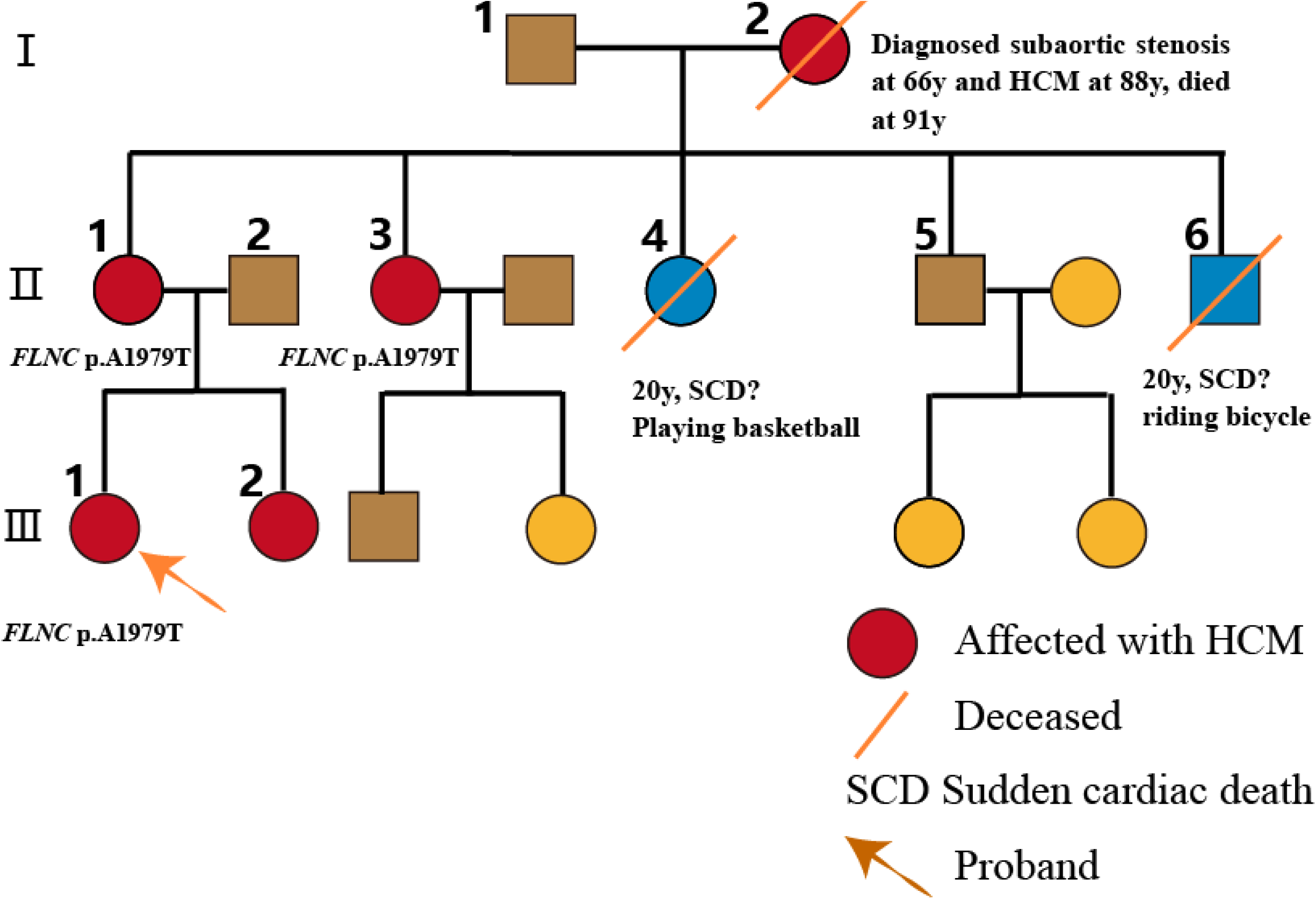
The pedigree chart. The proband and her family harbor the FLNC mutation and have a family history of sudden death. The proband (arrow) and her sister, her mother and her aunt all harbor the FLNC c.G5935A (p.A1979T) mutation (black boxes). Sudden death was observed in two members, who both died at the age of 20. It is not known whether the proband’s deceased relatives harbored the FLNC mutation.

### Searching for disease mutation(s)

We hypothesized that a gene mutation caused HCM in this family. To identify possible causative genetic variants, we collected peripheral blood leukocytes from five alive family members listed above (i.e. the proband, the father, the mother, the maternal aunt, and the maternal uncle) for the isolation of genomic DNA. We performed whole-exome sequencing at the Bohao Medical Laboratory in Shanghai, using the Illumina HiSeq 2500 instrument (San Diego, CA, USA) to generate paired-end reads of 150 bp, with an average coverage of 95 times. We analyzed and filtered exome results for all modes of inheritance to generate a list of candidate variants (including missense, nonsense, splice site variants, and indels). After removing common polymorphisms, a total of 241 variants, with the potential to affect protein coding, were identified in these subjects. Consequently, variants with a minor allele frequency of >0.01 in East Asian samples (EAS) of the 1000 Genomic Project database, the ESP6500, or the ExAC database, were removed. A total of 37 variants remained. We used different bioinformatic algorithms (SIFT(12), Polyphen2(13), FATHMM(14), MutationTaster(15), and CADD(16)) to search for variants affecting protein coding and genes that show cardiac-specific expression. Suspected causal variants were validated using Sanger sequencing.

### Directed mutagenesis of full-length Filamin C

We acquired a full-length cDNA clone for *FLNC*, with a C-terminal DYKDDDDK Tag, and a p.A1979T mutation clone in Vector GV362 from Shanghai Genechem Co., LTD.

### Cell culture and immunofluorescence

To evaluate whether the *FLNC* variant found in this family would impair the function of filamin C, full-length cDNA clones, which contained the p.A1979T mutation or the wild type, were generated and nucleofected into AC16 human cardiomyocytes. The cells were cultured in DMEM, which was supplemented with 10% fetal bovine serum (Life Technologies, USA). To express wild type and mutant filamin C, cells were transfected using X-tremeGENE Hp DNA Transfection Reagent (ROCHE, Switzerland), following the manufacturer’s instructions. For immunofluorescence analysis, cells were fixed in 4% paraformaldehyde solution, rinsed in phosphate-buffered saline (PBS) and permeabilized with 0.5% Triton X-100. Cells were incubated overnight at 4 with DYKDDDDK-Tag rabbit primary antibodies (Cell Signaling Technology, USA) and diluted in 10% fetal bovine serum in 1 × PBS. After washing with TBST, slides were incubated with 1:500 Alexa 555-conjugated Donkey anti-Rabbit IgG (H+L) Highly Cross-Adsorbed Secondary Antibody (ThermoFisher, USA), for one hour at room temperature. After washing, nuclei were counterstained with Hoechst 33342 (ROCHE, Switzerland) and slides were mounted in prolong gold antifade reagent (Invitrogen, USA). Micrographs were obtained using a laser scanning confocal microscope (LSCM) (Leica, SP5-FCS, Germany). Western blot analysis was performed to examine the expression level of the protein.

### Results

Among a total of 37 rare candidate variants with the potential to affect protein function, only one variant, *FLNC* p.A1979T (NM_001458: exon36: c. G5935A), showed co-segregation in affected family members, but absent in unaffected members, according to the sequencing of the five family members (□-1, □-1, □-2, □-3, and □-5). This variant was absent from the 1000 Genomes Project, the GO ESP database and the ExAC database. The variant was predicted to be damaging by SIFT, Polyphen2, and MutationTaster. Consequently, we validatied the variant using Sanger sequencing (Fig.2).

**Fig.2.**
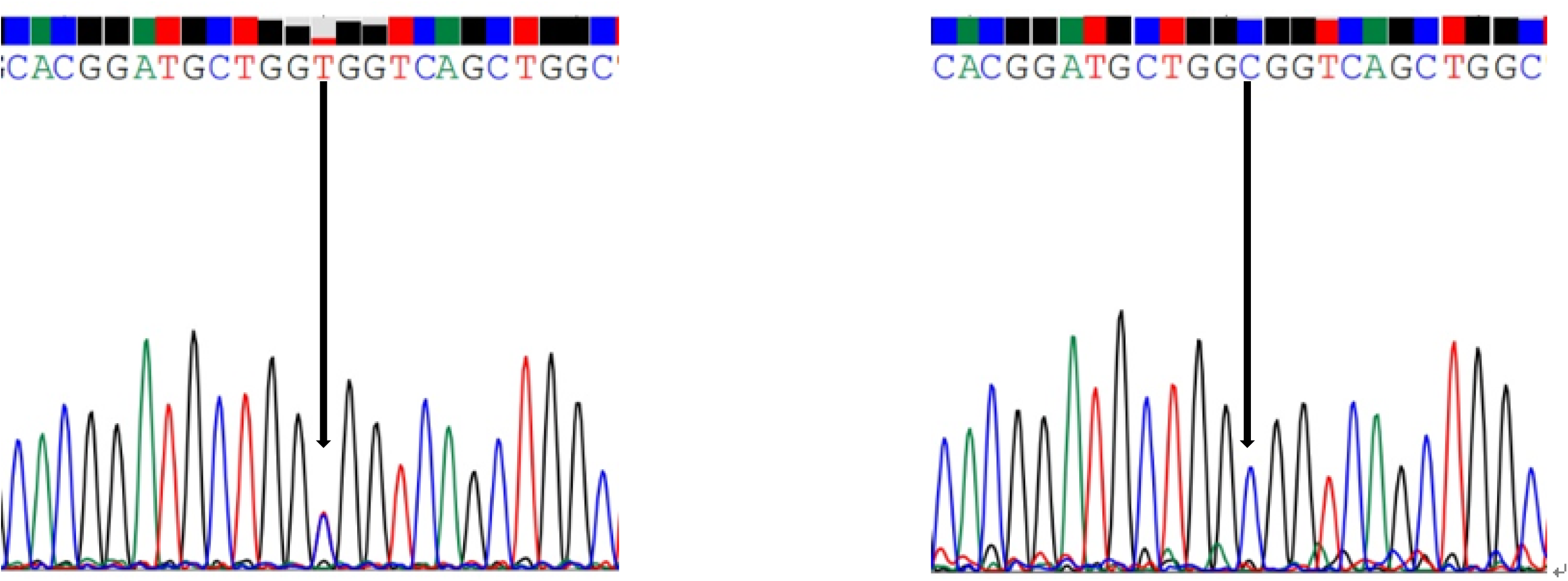
Results of Sanger sequencing of family member □-1 with HCM and family member □-5 without HCM in the family. The data show that the identified variant (p.A1979T) segregates with the disease.

The Gene Ontology (GO) annotation of *FLNC* and FLNC-related diseases (Fig.3A) with the online tool -ToppGene Suite (https://toppgene.cchmc.org/) showed that the function of *FLNC* was highly associated with skeletal muscle and cardiomyopathies. Multiple sequence alignments, including 100 vertebrates from the UCSC database (http://genome.ucsc.edu/), showed that the mutation site of *FLNC* (p.A1979T) is highly conserved (Fig. 3B). HOPE (Have (y)Our Protein Explained, http://www.cmbi.ru.nl/hope/) analysis schematically indicated structural change in a highly conserved positon resulting from substituting a Threonine for an Alanine at position 1979 in the *FLNC* protein (Fig.4A). This change resulted in a larger residue and increased hydrophobicity (17) (Fig.4B). The mutant residue maps to a highly conserved position, and lies in a domain, i.e. one of highly homologous immunoglobulin-like domains, which is important for binding with other molecules. Subcellular distribution assessed by immunofluorescence and confocal microscopy in our study showed that cells with wild type *FLNC* had a normal cytoplasmic distribution of the filamin C protein. However, cytoplasmic protein aggregations were present in cells carrying the p.A1979T mutant (Fig.5).

**Fig.3.**
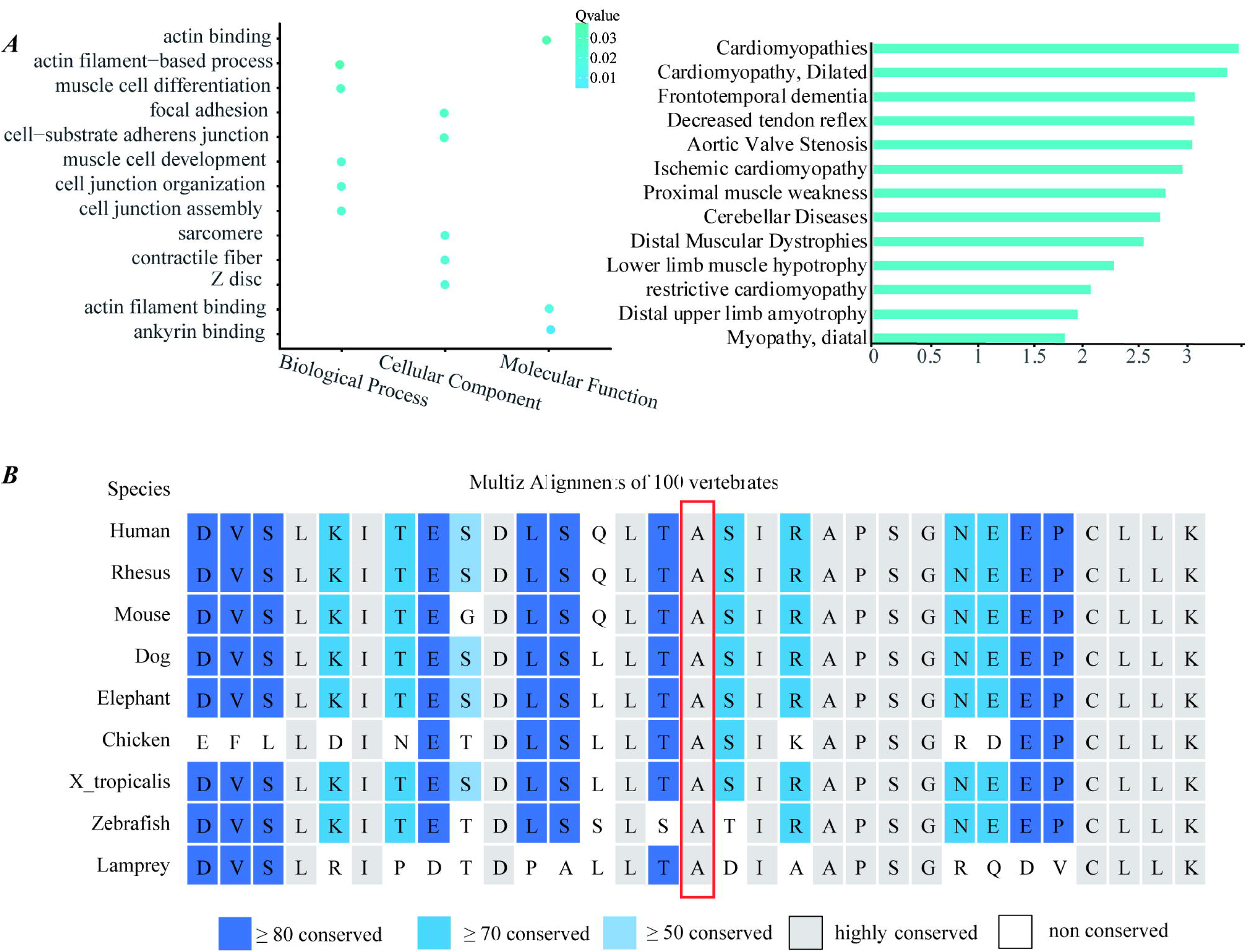
**(A)** Annotation of FLNC from ToppGene. **(B)**Amino acid conservation analysis in the mutant region. The mutant amino acid (A 1979) is in the red box.

**Fig. 4.**
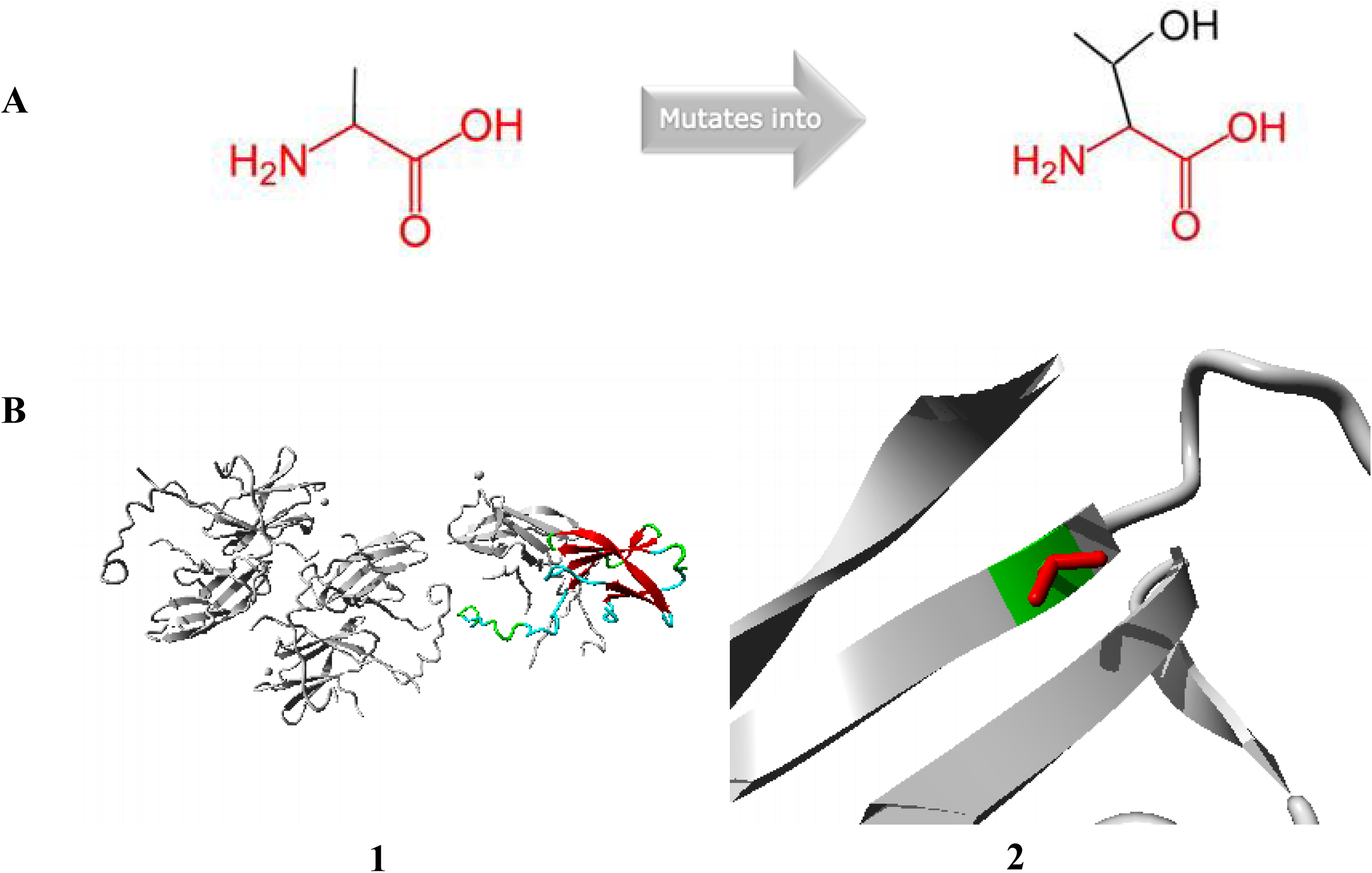
The results of HOPE (**A)** The chemical structural changes of the mutant amino acid. (**B1**) Overview of the protein in ribbon-presentation. The protein is coloured by element; helix= blue; strand = red; turn = green, 3/10 helix = yellow and random coil = cyan. Other molecules in the complex are colored grey when present. (**B2**) Close-up of the mutation. The protein is colored grey, the side chains of both the wild-type and the mutant residue are shown and colored green and red respectively.

**Fig.5.**
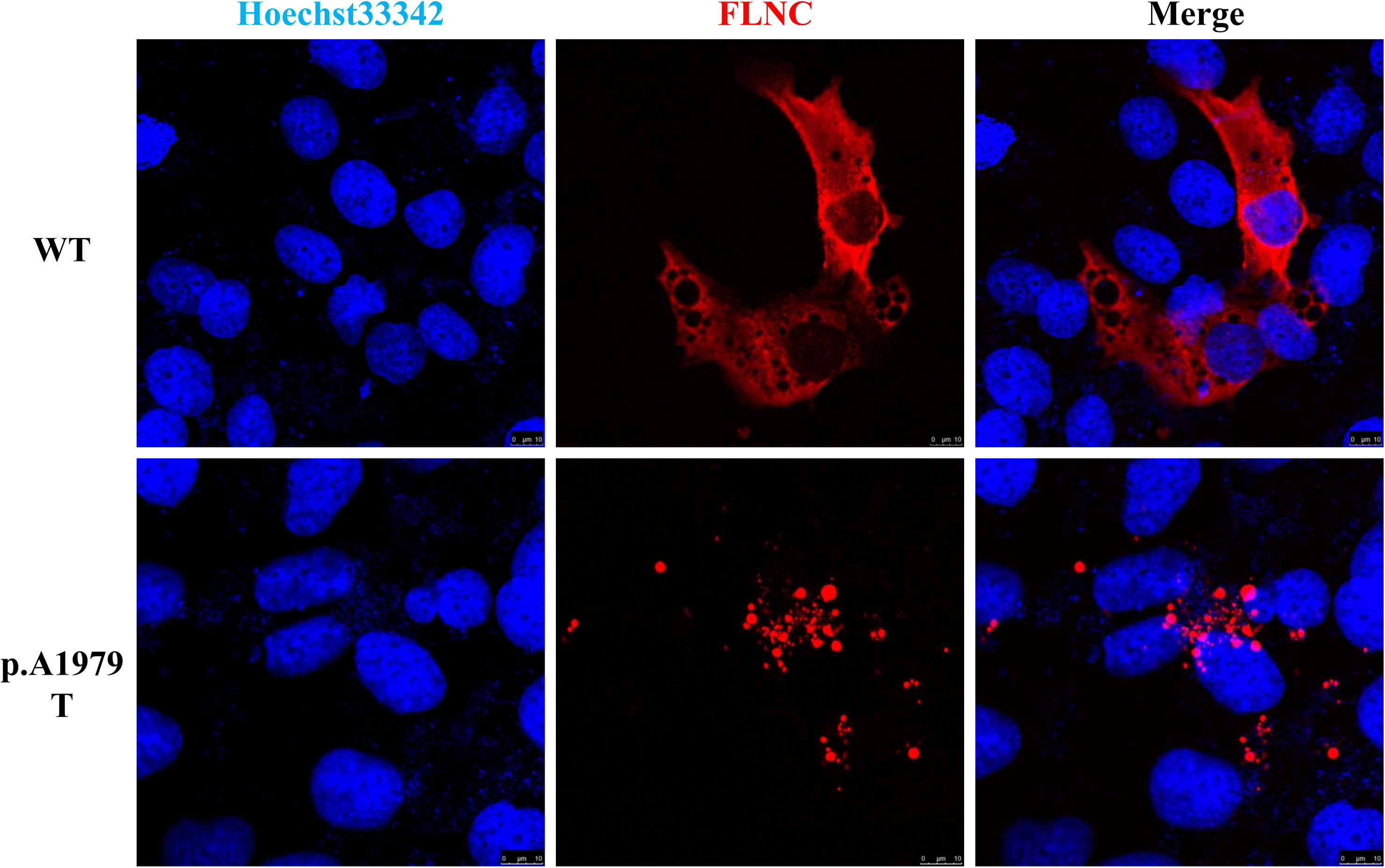
Function analysis of the *FLNC* mutant. The AC16 cells were transiently transfected with expression vectors encoding DYKDDDDK-tag (DDK-tag) wild type (WT) *FLNC* or mutant p. A1979T. Subcellular distribution of ectopically expressed WT and mutant *FLNC* in AC16 cells was detected by immunofluorescence and confocal microscopy. The filamin C distribution was detected by anti-DDK tag antibody and nuclei were counterstained with Hoechst33342.

Interestingly, a second mutation, NM_033118:exon8:c.G1138T:p.E380X of the *MYLK2* gene, was identified in the mother (□-1) of the proband, but absent in other subjects. This truncating mutation was predicted as damaging by SIFT. A number of studies have reported *MYLK2* muations causing HCM(18, 19). In addition, the study by Davis et al. suggested that *MYLK2* muations may serve as the cause of a digenic form of HCM(20). Different from the previous report and distinct from other affected family members, the case □-1 as a compound hetezygote of *FLNC* p.A1979T and *MYLK2* p.E380X has a clinical diagnosis of DCM. Truncated transcripts and related nonsense-mediated decay from this mutation may help to explain the atypical clinical features of □-1, including her dilated cardiomyopathy and heart failure.

## Discussion

*FLNC* mutations were initially recognized to be related to a specific form of skeletal MFM, combined with an unspecified type of “cardiomyopathy” (7, 9, 21, 22). In a recent study, Juan Gómez et al. reported six likely pathogenic variants in seven (1.5%) patients out of 448 HCM patients in Spain, and concluded that most of the *FLNC* variants were associated with mild forms of HCM and reduced penetrance (23). Since 2012, patients with inherited cardiomyopathy and the incidence of SCD take genetic screening including the *FLNC* gene(6). The nonsynonymous A1979T mutation identified in this study mapps to one of rod domains of filamin C, and a number of mutations in these domains have been identified of causing familial HCM(5).

*FLNC* mutations can cause different cardiomyopathies (e.g. dilated, hypertrophic, restrictive, and arrythmogenic cardiomyopathy), but the association between the mutation type and specific cardiac phenotype is elusive. While *FLNC* missense mutations may be associated with HCM or RCM, nonsense mutations leading to a truncated protein typically cause DCM and/or arrhythmias (6, 23-26). Furst et al. reported that missense mutations could lead to changes in *FLNC* function, and truncated transcripts often undergo nonsense-mediated decay, which results in haploinsufficiency (27). Thus in the case of *FLNC* truncations, haploinsufficiency may be the cause of both arrhythmias and DCM (6), while missense mutations can damage the protein function and result in HCM and RCM.

What intriguted us to do the functional experiment on the novel mutation *FLNC* p.A1979T was two different phenotypes were seen in this family (DCM in □-1 and HCM in other affected family members), with the purpose to exclude complex effects on the *FLNC* gene (e.g. both protein level and structure). For the functional experiment, while rat or human myoblast cell models were used in previous reports, our study utilized human cardiomyocytes (AC16) to represent better *in vivo* effects of the mutation. As shown in our experiments, the *FLNC* p.A1979T mutation in human cardiomyocytes can cause cytoplasmic aggregations and impair protein function, in concordant with the previous studies with other cell models(5, 23). At the same time, our western blot analysis showed that *FLNC* is expressed equivalently in both wild type and mutant. Instead, cytoplasmic protein aggregations were resulted by the mutation.

In many cases, both arrhythmias and cardiomyopathies are frequently present in the same patient, which is most likely due to the causative effect of one on the other. The family in this study, which has an *FLNC* missense mutation, has an almost normal rhythm, which is in line with other reports. Cases of HCM with *FLNC* mutations have a higher proportion of sudden cardiac deaths when compared with cases without *FLNC* mutations (5). In the family studied here, the proband’s maternal uncle and maternal aunt both died of SCD during physical exercise at 20 years old.

Ortiz-Genga et al. reported the association of truncating mutations in *FLNC* with an overlapping phenotype of DCM and left-dominant arrhythmogenic cardiomyopathies, complicated by frequent, premature, SCD (6). *FLNC* is a part of membrane proteins in the actin network. *FLNC* maybe coordinates the activation of ion channels in cell membranes, and loss-of-function leads to cardiac conduction disorders. Neethling et al. reported that KCNE2, which is a voltage-gated potassium channel and functions in ventricular repolarization, interacted with *FLNC* (28). Given the collective evidence, the SCD in this family might result from incidental malignant arrhythmias. However, *FLNC* p.A1979T can’t explain the DCM of the mother.

Instead, the compound effect of a second truncating mutation, p.E380X of the *MYLK2* gene, in the mother (□-1) of the proband, suggested a digenic form of cardiomyopathy. Highlighted by our study, the possibility of a second causal mutation of cardiomyopathy warrants clinical attention, especially for patients with atypical clinical features.

## Data Availability

The dataset generated for this manuscript is the whole exome sequencing data and not publicly available by the national laws of China.

## Author Contributions Statement

XQ and PL: performing the cell experiments, design of the study, analysis and interpretation of data, critical revision of the manuscript;

YX, SC, HQ, YL, LT: analysis and interpretation of data, drafting manuscript, critical revision of the manuscript content;

YY, FH and PW: acquisition of clinical data, analysis of data, critical revision of the content

XZ: acquisition of clinical data of patients;

XL, YW, and RZ: design of the study;

YW, HH, and JZ: design of the study, handling funding, writing the paper, full text evaluation and guidance, final approval of the version to be submitted.

All authors read and approved the final manuscript.

## Declarations

Ethics approval and consent to participate. Clinical tissue specimens were obtained from the Guangdong Provincial People’s Hospital. This study was approved by the ethics committee of the Guangdong Provincial People’s Hospital (Grant No. GDREC2016255H).

## Acknowledgments

The authors would like to thank the family for their participation.

